# A comprehensive evaluation of minimally invasive Achilles tendon reconstruction with hamstring graft indicates satisfactory long-term outcomes

**DOI:** 10.1101/2022.08.02.22278266

**Authors:** Bartosz Kiedrowski, Paweł Bąkowski, Paweł Cisowski, Łukasz Stołowski, Jakub Kaszyński, Magdalena Małecka, Tomasz Piontek

**Affiliations:** Department of Orthopedic Surgery, Rehasport Clinic, Poznań, Poland; Sport Championship School on the Handball Federation in Poland, Płock, Poland; Department of Spine Disorders and Pediatric Orthopedics, University of Medical Sciences Poznań, Poznań, Poland

**Keywords:** Achilles tendon tears, minimally invasive Achilles tendon reconstruction, a comprehensive evaluation of Achilles tendon

## Abstract

Given the relevance of the Achilles tendon in proper function of the foot and ankle, the primary goal of the present study was to use a holistic approach for a comprehensive evaluation of Achilles tendon reconstruction results on multiple levels. 30 patients with partial or total Achilles tendon tears were subjected to the minimally invasive Achilles tendon reconstruction. Patients were then subjected to the clinical, functional and isokinetic tests 12 and 24 months after the treatment. The overall results of this extensive evaluation are highly satisfactory and patients returned to their normal physical activity.

## Introduction

The Achilles tendon is the largest and strongest tendon in the body. A partial or a total rupture of this tendon can occur with a sudden strain or a stretch. The incidence of Achilles tendon ruptures is 18 - 40 per 100 000 persons per year and occurs more frequently in the fourth decade of life [1,2]. The repair of the ruptured Achilles tendon is critical for proper function of the lower limb. For years, the Achilles tendon has been repaired through a large open incision. This technique required full tendon exposure and the repair was done in the damaged area of the tendon. Recently, percutaneous, minimally invasive repair techniques are getting more interest, since the surgery is done through small incisions and the stitches are placed in the strong, healthy area of the tendon. Moreover, the tendon exposure is no longer necessary and this decreases a scar formation and a wound healing issues and allows more rapid recovery, earlier physical therapy and quicker return to the activity. However, in case of Achilles tendon re-ruptures, degeneration or chronic tears, when the tendon tissue is damaged, a reconstruction technique using a graft is needed.

Despite the surgical method used, the outcomes of the Achilles tendon repair have to be certainly evaluated. Multiple clinical and functional examination procedures have been designed for that purpose. This includes patient-reported subjective tests (Hannover Achilles Score, Leppilahti Score, Rupp Achilles Tendon Score, Achilles Tendon Total Rupture Score, American Orthopedic Foot and Ankle Society Score and more [3]) and objective clinical evaluation by a physiotherapist or an orthopedist. Moreover, the characteristics of the skeletal muscles under dynamic conditions can be objectively measured using specialized devices, isokinetic dynamometers. However, it has been recently recognized that there is no consensus on the best or most appropriate way to measure the strength after Achilles tendon reconstruction, irrespective of a treatment [4]. No unified protocol exists for establishing strength after Achilles tendon reconstruction.

In our recent research we have shown that a minimally invasive, endoscopic Achilles tendon reconstruction using semitendinosus and gracilis tendons with Endobutton stabilization allows for good functional recovery [5,6]. As a logical consequence, the primary goal of the present study was to use a holistic approach for a comprehensive evaluation of Achilles tendon reconstruction results on multiple levels: clinical, functional and isokinetic. Given the relevance of the Achilles tendon in proper function of the foot and ankle, we reasoned that such exhaustive evaluation of the recovery of motor function requires not only a clinical or subjective assessment, but also a quantitative analysis of muscle strength and endurance. To our knowledge, this is the first report aimed at revealing the results of both, subjective and objective tests performed in isolated and functional conditions. Moreover, we have separately evaluated the results obtained by patients with a partial and a total Achilles tendon tears.

## Materials and Methods

### Patients characteristic and treatment

30 patients with Achilles tendon tears were included in the study. All patients were treated surgically with a minimally invasive Achilles tendon reconstruction technique using a hamstring autografts, as previously described [5]. The surgical procedures were performed by two trained orthopedic surgeons (PB and TP) between 2016 and 2019 at Rehasport Clinic in Poznań. All patients were followed the proprietary rehabilitation protocol developed at our Clinic [7].

The whole group of 30 patients was examined 12 months after the procedure. At this time-point we have distinguished and evaluated separately two subgroups: (i) seventeen patients, which suffered from a partial tendon rupture, and (iii) thirteen patients, which had a total tendon injury. 21 patients were additionally evaluated 24 months after the procedure, however due to the lower subgroup’s sizes (12 patients with partial and 6 with total tendon rupture), the evaluation was performed for the whole group solely. The demographic data of the patients enrolled in the study are presented in Table 1.

**Table 1.**
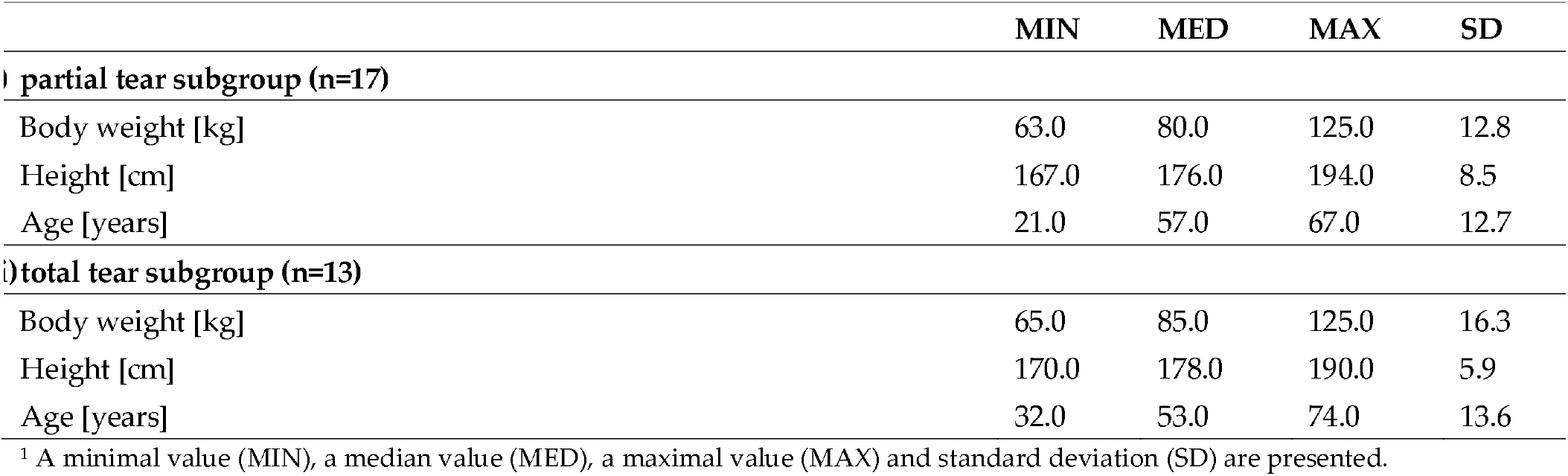
Demographic data of the patients enrolled in the study.

### Clinical and subjective evaluations

Clinical examination was performed by trained physiotherapist (BK) and included calf circumference measurements. Additionally, the subjective outcomes of the surgery reported by patients were recorded, as previously described. These tests were based on the pain and the satisfaction levels and included the visual analogue scale (VAS scale), the Achilles tendon Total Rupture Score (ATRS) [8] and the quality of life questionnaire (EQ-5D-5L) [9].

### Functional evaluations

The functional evaluation was performed by trained physiotherapist (BK) and was based on three tests: the weight bearing lunge test, the heel rise test and single leg hop. All tests were performed for each limb separately, following the existing protocols [10]. The weight bearing lunge test was performed to measure the extent of dorsiflexion of the ankle joint and foot complex [11]. The heel rise test was performed to measure the endurance of the Achilles [12]. A single leg hop test was used for measuring the distance of the horizontal jump [13].

### Isometric and isokinetic evaluation

Isometric and isokinetic evaluation was performed using Biodex 3 dynamometer as previously descried [14]. Both, the injured and non-injured legs were tested to assess an isometric, concentric and eccentric muscle strength parameters. An average isokinetic knee extension and flexion peak torques were measured at concentric and eccentric velocity of 30°/s. The total contractional work, the average power (the average time rate of work) of the muscles as well as the maximum peak torque (angle peak torque) was appointed at 90°/s velocity. The explosive power of the muscles (time to peak torque) was evaluated at three concentric and eccentric velocities: 30°/s, 60°/s, and 90°/s. Testing started with the 30°/s angular velocity and then increased to higher velocities. An average of three repetitions was performed for each angular velocity for both affected and intact limbs. Before testing, practice trials were given to the patients to familiarize them with the machine.

### Statistical analysis

The sample size calculation showed that with a power of 80% (2-sided testing at a significance level of 0.05), a sample size of 11 participants was needed to show a difference between operated and non-operated limb and between partial and total Achilles tear subgroups [15]. The statistical analysis of the results has been carried out in the R environment. For all measurements, statistical significance was considered for p <0.05. Statistical characteristics of individual measurements has been carried out through the analysis of the distribution of values. Pearson’s correlation tests between parameters were performed.

## Results

### Clinical and subjective evaluations

The results of the calf circumference examination are presented in Table 2. We have observed a statistically significant difference (p=0.0479) in the circumference of the operated vs non-operated limb 12 months after the surgical procedure (Fig. 1A). This difference was observed in the total Achilles rupture subgroup solely (Fig. 1B, p=0.0161). There was no statistically significant difference in circumference between the operated and non-operated limbs after 24 months. The injured leg showed slightly increased maximum calf circumference than the opposite leg in almost all cases, but the difference was not significant.

**Table 2.**
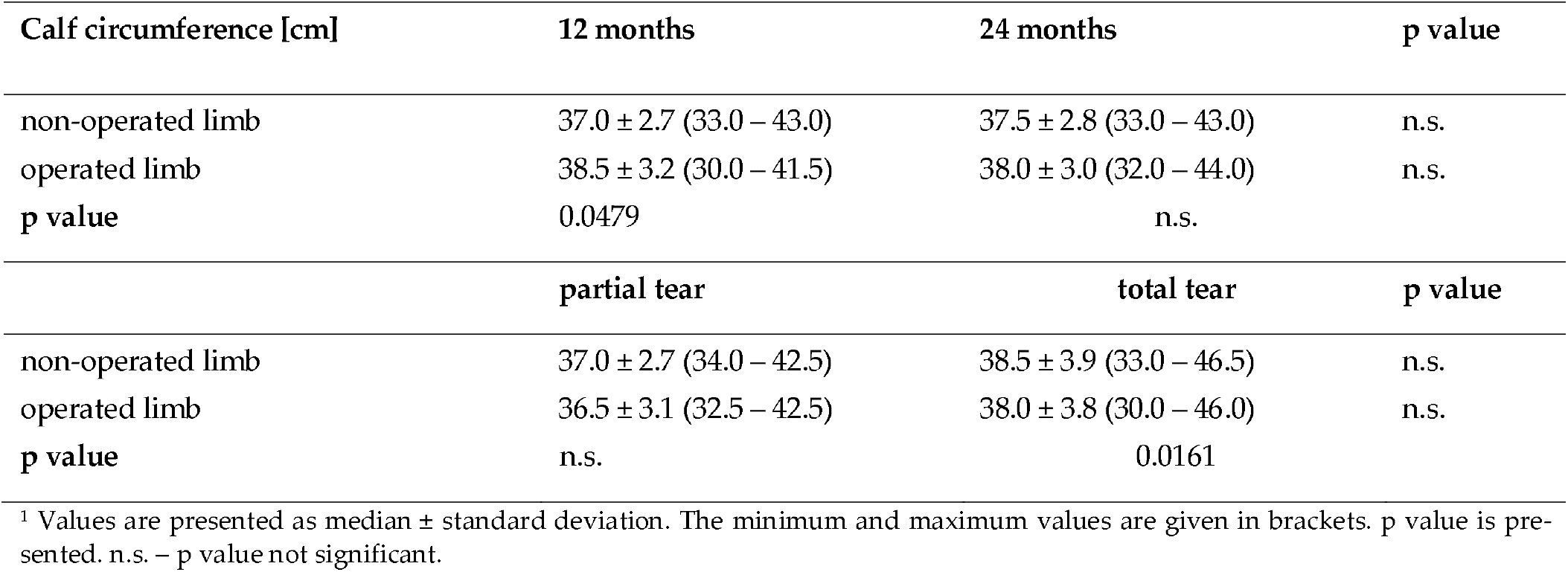
Clinical evaluation of calf circumference.

**Figure 1.**
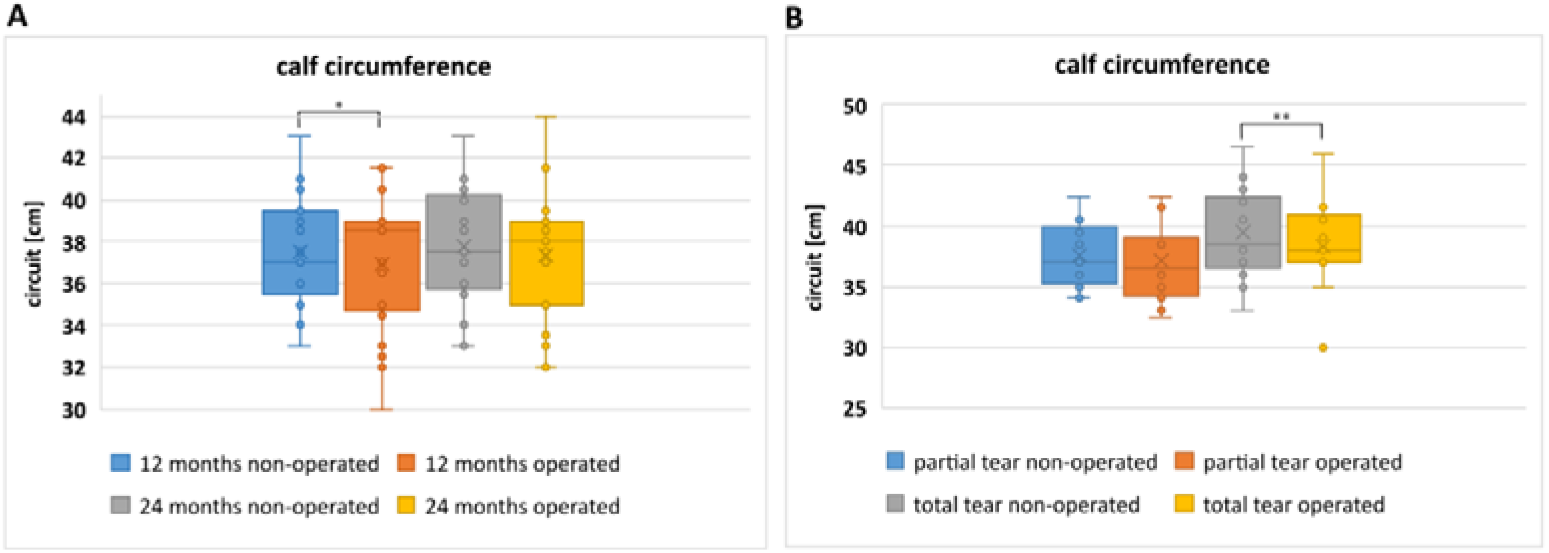
Clinical evaluation of calf circumference differences between (**A**) two evaluations appointed at 12 and 24 months postoperatively; (**B**) two subgroups of partial and total Achilles tendon tear.

The subjective outcomes reported by patients 12 and 24 months after the surgical treatment are gathered in Table 3. The results obtained in the subjective tests did not differ statistically between the partial and the total Achilles tear subgroups (Fig. 2B). The ATRS score improved significantly 24 months after the surgery (p=0.009, Fig. 2A), with 4 patients reaching the maximum score of 100 points. The median value of the EQ-5D-5L scale improved statistically (p=0.0087), as well as EQ-5D-5L health comfort test increased significantly, from 75.0 to 90.0 (p=0.0008). The median reported pain and satisfaction level according to the VAS scale was 1.0 (±1.5) and 9.0 (±1.4) 12 months after the procedure, respectively, and 0.0 (±0.9) and 10.0 (±0.4) at the second evaluation (p=0.007 and p=0.0143, respectively). The maximum level of VAS satisfaction score (10 points) was recorded by 10 participant after 12 months postoperatively and 13 at 24 months follow-up.

**Table 3.** The subjective outcomes of the surgical procedure.

| Test | 12 months | 24 months | p value |
| --- | --- | --- | --- |
| ATRS | 79.5 ± 12.7 (58.0 – 99.0) | 90.0 ± 9.5 (72.0 – 100.0) | 0.0009 |
| EQ-5D-5L | 6.0 ± 2.6 (5.0 – 14.0) | 6.0 ± 2.0 (5.0 – 13.0) | 0.0097 |
| EQ-5D-5L Health today | 75.0 ± 15.9 (30.0 – 100.0) | 90.0 ± 14.1 (40.0 – 95.0) | 0.0008 |
| VAS pain | 1.0 ± 1.5 (0.0 – 5.0) | 0.0 ± 0.9 (0.0 – 3.0) | 0.0007 |
| VAS satisfaction | 9.0 ± 1.4 (5.0 – 10.0) | 10.0 ± 0.7 (8.0 – 10.0) | 0.0143 |
|  | partial tear | total tear | p value |
| ATRS | 84.0 ± 14.4 (50.0 – 99.0) | 86.5 ± 24.8 (7.0 – 96.0) | n.s |
| EQ-5D-5L | 6.0 ± 2.2 (5.0 – 14.0) | 6.0 ± 1.2 (5.0 – 9.0) | n.s |
| EQ-5D-5L Health today | 80.0 ± 16.4 (30.0 – 100.0) | 80.0 ± 11.9 (65.0 – 100.0) | n.s |
| VAS pain | 1.0 ± 1.7 (0.0 – 5.0) | 0.0 ± 1.0 (0.0 – 3.0) | n.s. |
| VAS satisfaction | 9.0 ± 1.1 (7.0 – 10.0) | 10.0 ± 1.1 (7.0 – 10.0) | n.s. |

<sup>1</sup> Values are presented as median ± standard deviation. The minimum and maximum values are given in brackets. p value is presented. n.s. – p value not significant.

**Figure 2.**
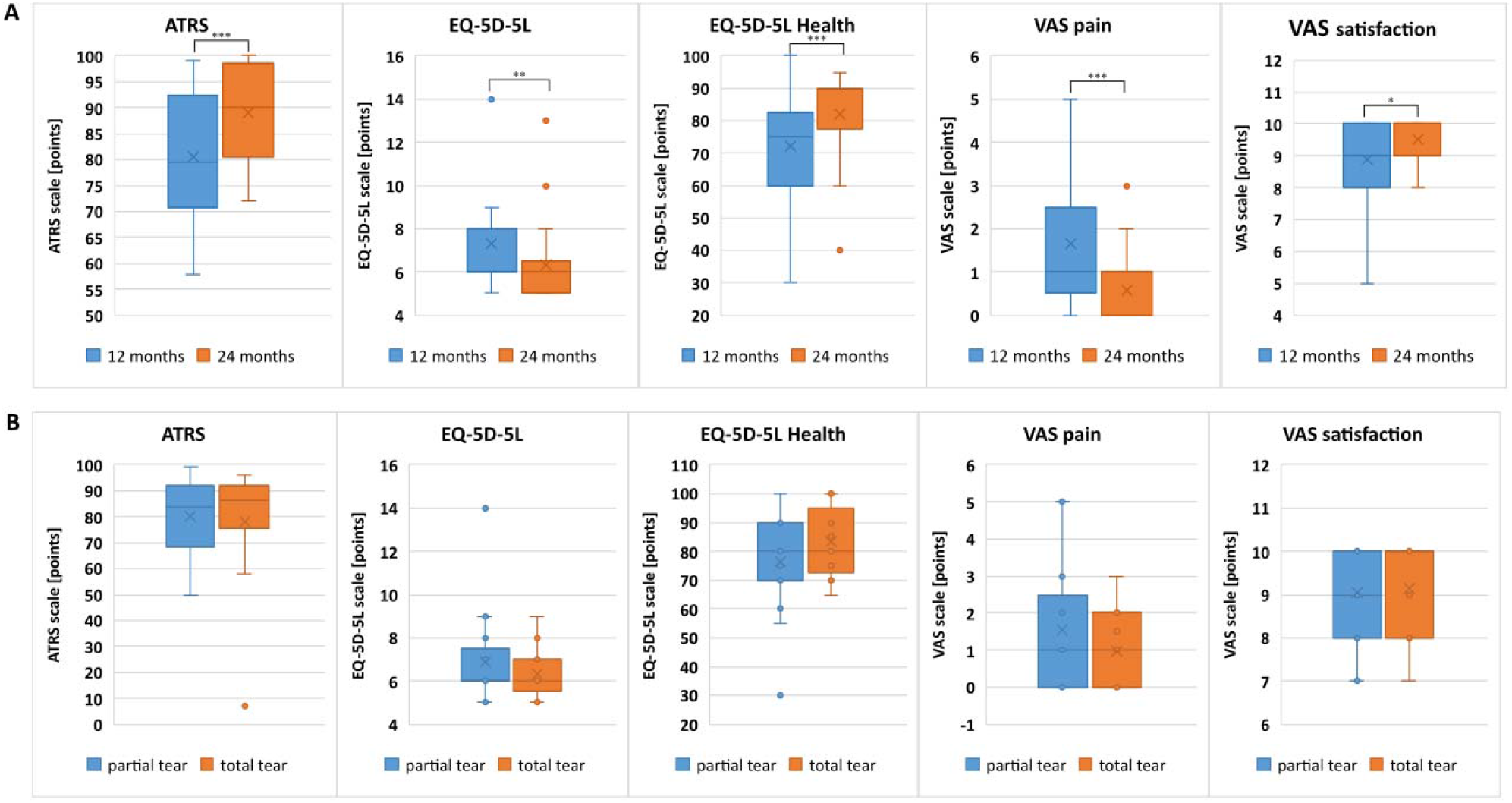
Evaluation of patient-reported subjective differences between operated and non-operated limbs (**A**) at 12 and 24 months postoperatively; (**B**) in two subgroups of partial and total Achilles tendon tear.

### Functional evaluation

We have observed statistically significant differences in performing all functional tests with an injured extremity between the two follow-ups (Table 4). The results of the functional evaluation of non-operated limbs did not change significantly betweeen 12 months and 24 months after the surgery (Fig. 3A). Generally, significantly better results in the function of the operated limbs were obtained 24 month after the surgery. However, a significant difference was still observed in the functional tests at both follow-ups, when we compared the operated vs non-operated limb (with a fovour of non-operated one), both in the partial and the total tear subgroups (Fig. 3B). Patients were able to reach the larger distance from the wall to the tip of the big toe at 24 months follow-up (when compared to 12 months, p=0.0118) during the weight bearing lunge test with the operated limb. Smiliar situation was observed in the number of repetitions of the heel rise in the heel rise test (p=0.0001). The largest difference in number of repetitions performed with the operated limb at 12 months after the surgery was 14 (19 for the uninjured limb and 5 for the injured limb in two cases). 24 months after the surgery we observed clearly smaller differences: the same number of heel rise repetitions was observed for non-operated and operated limb in one patient and a difference of one heel rise – in 6 cases. The differences in the distance of the injured (p=0.0037) and non-injured (p=0.0253) leg hop were statistically significant when we compared 12 months vs 24 months postoperatively, with better results obtained at the second evaluation.

**Table 4.** The functional outcomes of the surgical procedure.

|  | **12 months** | **24 months** | **p value** |
| --- | --- | --- | --- |
| **weight bearing lunge test** |  |  |  |
| non-operated limb | 13.0 ± 2.9 (5.0 – 18.0) | 12.0 ± 2.0 (5.0 – 19.0) | n.s. |
| operated limb | 10.0 ± 2.8 (2.0 – 14.0) | 10.0 ± 2.5 (5.0 – 15.0) | 0.0118 |
| **p value** | 0.0002 | 0.0030 |  |
| **heel rise test** |  |  |  |
| non-operated limb | 16.0 ± 6.2 (2.0 – 22.0) | 16.0 ± 5.4 (3.0 – 26.0) | n.s. |
| operated limb | 9.0 ± 6.0 (0.0 – 21.0) | 13.0 ± 5.4 (3.0 – 22.0) | 0.0001 |
| **p value** | 0.0000 | 0.0000 |  |
| **single leg hop** |  |  |  |
| non-operated limb | 100.0 ± 27.5 (40.0 – 140.0) | 110.0 ± 34.0 (45.0 – 190.0) | 0.0235 |
| operated limb | 79.0 ± 29.7 (40.0 – 145.0) | 105.0 ± 35.7 (40.0 – 180.0) | 0.0037 |
| **p value** | 0.0010 | 0.0063 |  |
|  | **partial tear** | **total tear** | **p value** |
| **weight bearing lunge test** |  |  |  |
| non-operated limb | 12.0 ± 2.7 (5.0 – 16.0) | 12.0 ± 3.2 (3.0 – 16.0) | n.s. |
| operated limb | 10.0 ± 2.4 (4.0 – 13.0) | 7.0 ± 3.5 (2.0 – 14.0) | n.s. |
| **p value** | 0.0064 | 0.0063 |  |
| **heel rise test** |  |  |  |
| non-operated limb | 16.0 ± 6.1 (2.0 – 22.0) | 17.0 ± 5.8 (4.0 – 22.0) | n.s. |
| operated limb | 10.0 ± 6.5 (0.0 – 21.0) | 9.0 ± 5.4 (0.0 – 15.0) | n.s. |
| **p value** | 0.0000 | 0.0008 |  |
| **single leg hop** |  |  |  |
| non-operated limb | 97.5 ± 36.0 (40.0 – 175.0) | 100.0 ± 35.8 (45.0 – 150.0) | n.s. |
| operated limb | 82.5 ± 36.4 (40.0 – 165.0) | 178.0 ± 33.6 (45.0 – 140.0) | n.s. |
| **p value** | 0.0066 | 0.0013 |  |

<sup>1</sup> Values are presented as median ± standard deviation. The minimum and maximum values are given in brackets. p value is presented. n.s. – p value not significant.

**Figure 3.**
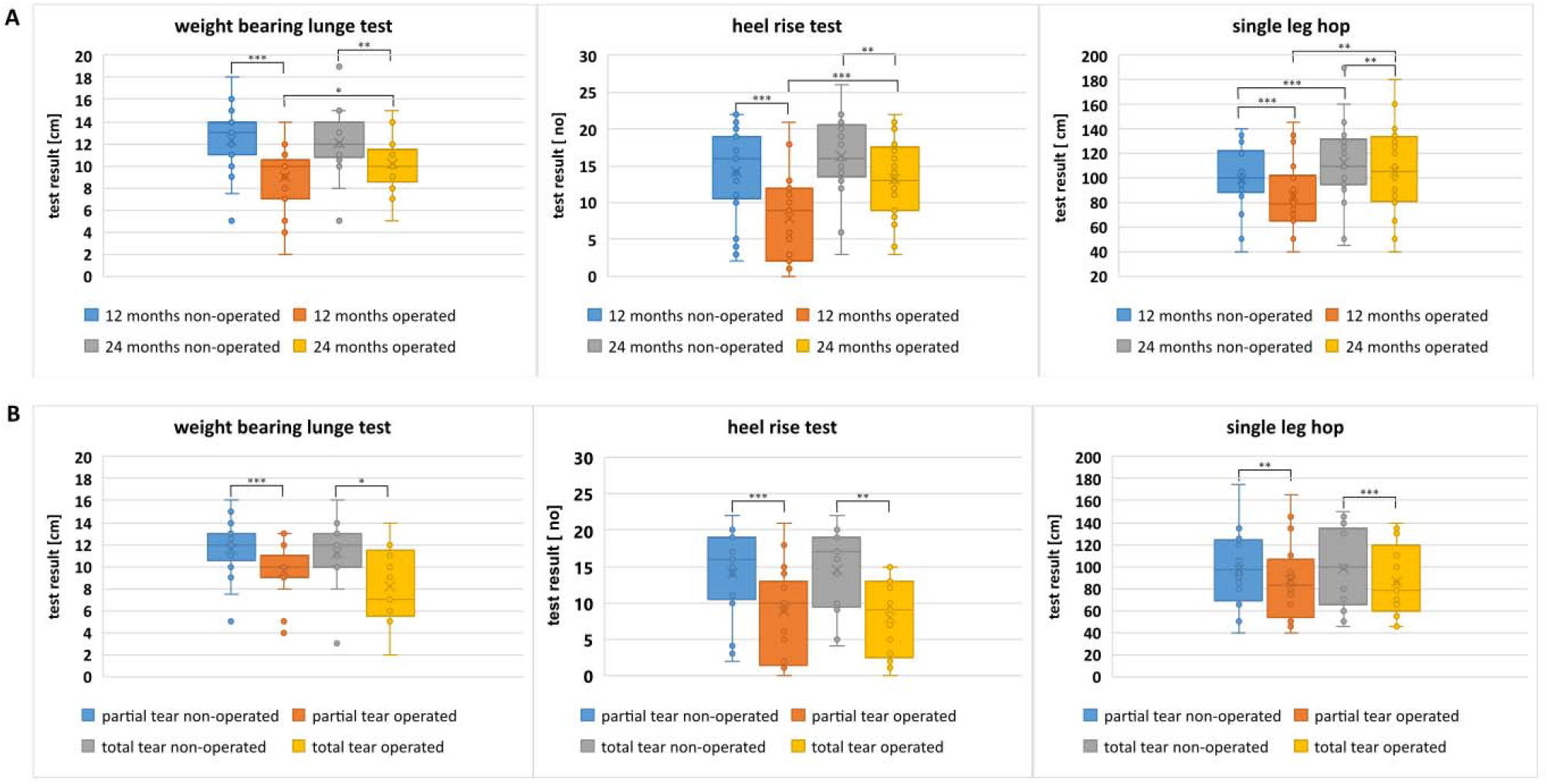
Evaluation of functional differences between operated and non-operated limbs (**A**) at 12 and 24 months postoperatively; (**B**) in two subgroups of partial and total Achilles tendon tear.

### Isometric evaluation

Lower limb strength is dependent on the body weight. Therefore, the limb strength was measured as an avarage peak torque (the turning effect of a given force on an object) in two motions: away from and towards the dynamometer, and expressed in relation to the patient body weight, in order to compare the results between individuals (Table 5). A statistically significant improvement was observed for the strenght of the operated limb, measured in “away” motion between the first and the second evaluation (p= 0.0364, Fig. 4A). The average peak torque/body weight in “toward” motion was comparable for operated and non-operated limbs in partial tear and total tear subgroups (Fig. 4B).

**Table 5.** The isometric outcomes of the surgical procedure.

|  | 12 months | 24 months | p value |
| --- | --- | --- | --- |
| **average peak torque/body weight away (%)** |  |  |  |
| non-operated limb | $1.2 \pm 0.3$ (0.4 – 1.6) | $1.1 \pm 0.3$ (0.6 – 1.9) | n.s. |
| operated limb | $0.8 \pm 0.4$ (0.3 – 1.7) | $1.0 \pm 0.3$ (0.4 – 1.6) | 0.0364 |
| **p value** | 0.0001 | 0.0008 |  |
| **average peak torque/body weight toward (%)** |  |  |  |
| non-operated limb | $0.6 \pm 0.1$ (0.4 – 0.7) | $0.6 \pm 0.2$ (0.3 – 1.1) | n.s. |
| operated limb | $0.6 \pm 0.2$ (0.1 – 0.8) | $0.6 \pm 0.2$ (0.4 – 1.0) | n.s. |
| **p value** | n.s. | n.s. |  |
|  | **partial tear** | **total tear** | **p value** |
| **average peak torque/body weight away (%)** |  |  |  |
| non-operated limb | $1.1 \pm 0.3$ (0.4 – 1.5) | $1.2 \pm 0.3$ (0.7 – 1.6) | 0.0174 |
| operated limb | $0.8 \pm 0.4$ (0.3 – 1.7) | $0.9 \pm 0.4$ (0.1 – 1.6) | n.s. |
| **p value** | 0.0327 | 0.0041 |  |
| **average peak torque/body weight toward (%)** |  |  |  |
| non-operated limb | $0.6 \pm 0.1$ (0.3 – 0.7) | $0.6 \pm 0.2$ (0.4 – 0.7) | n.s. |
| operated limb | $0.6 \pm 0.1$ (0.2 – 0.8) | $0.6 \pm 0.1$ (0.4 – 0.7) | n.s. |
| **p value** | n.s. | n.s. |  |

<sup>1</sup> Values are presented as median ± standard deviation. The minimum and maximum values are given in brackets. p value is presented. n.s. – p value not significant.

**Table 6.**
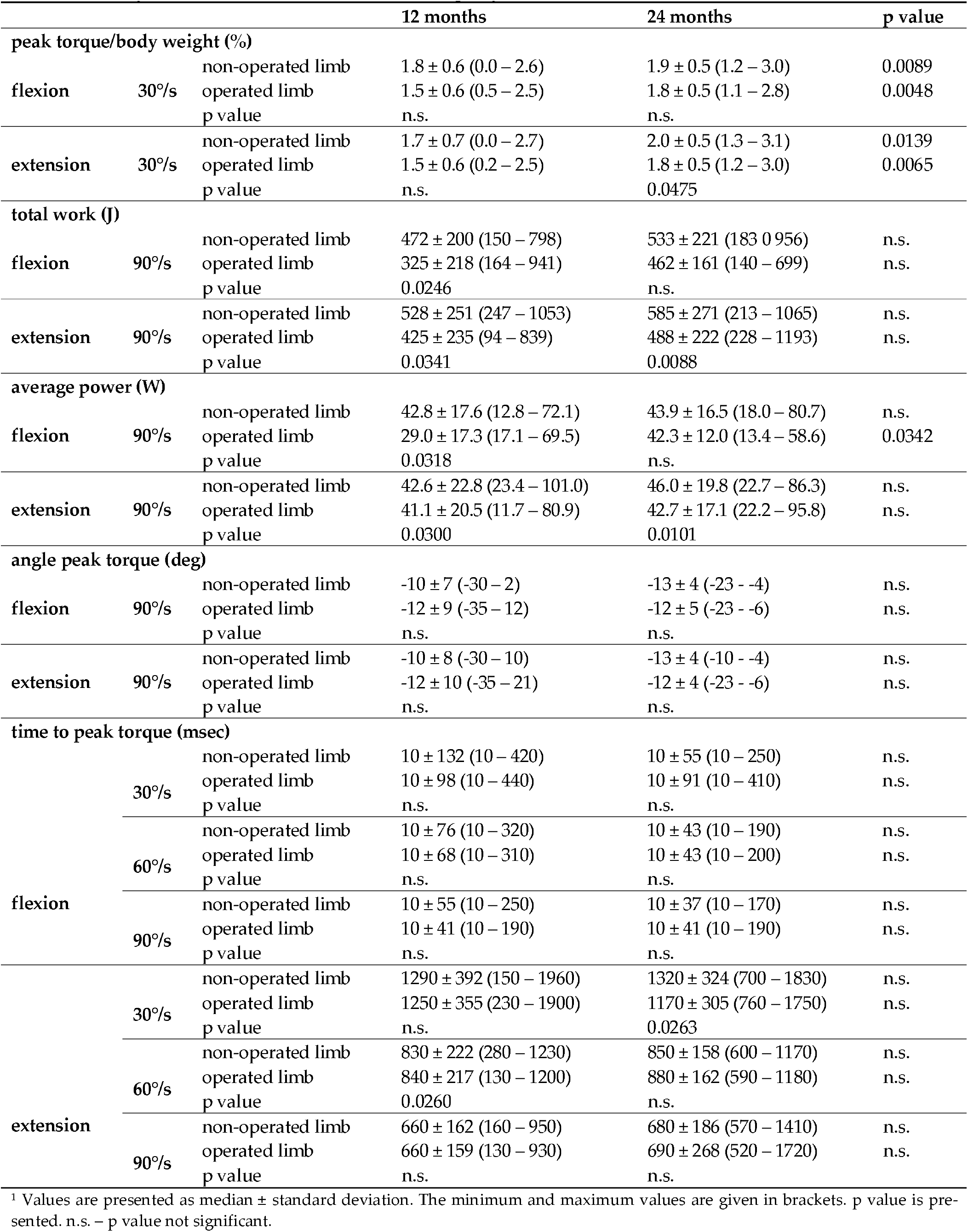
The comparison of the isokinetic outcomes of the surgical procedure at two evaluation times.

**Figure 4.**
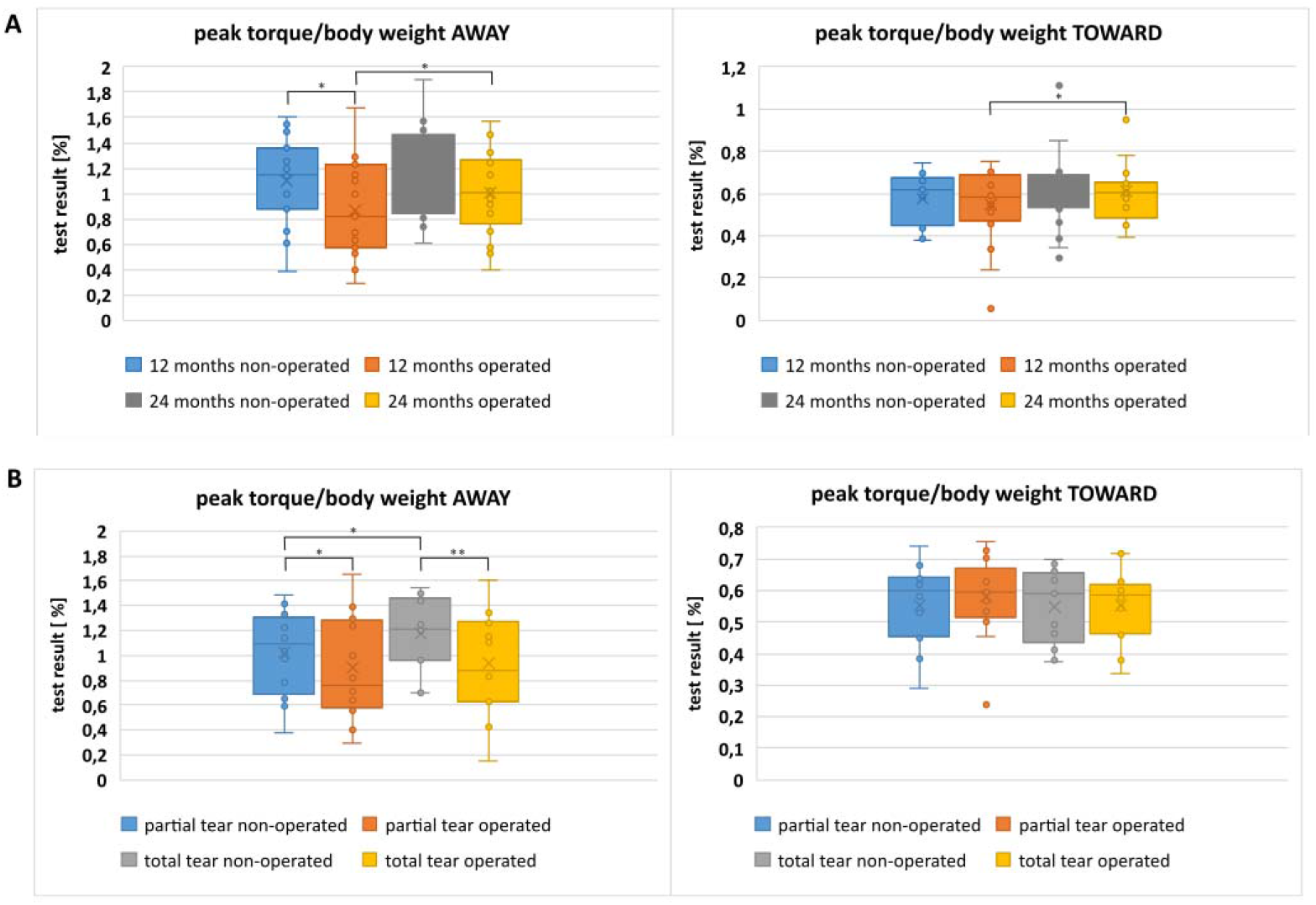
Evaluation of isometric differences between operated and non-operated limbs (**A**) at 12 and 24 months postoperatively; (**B**) in two subgroups of partial and total Achilles tendon tear.

### Isokinetic evaluation

Most of the musle strength parameters were comparable between two evaluation times (Table 5, p value n.s.). A statistically significant improvement was observed both, during the knee flexion and extension in operated limb, between the first and the second evaluation in the peak torque/body weight parameter (p= 0.0048 and 0.0065, respectively). Similar results were also noticed for non-operated limb (Fig. 5A). The average power of the muscles in the operated extremities significantly improved through the course of evaluations (p=0.0342). However, the average muscle power during the knee extension did not improve and the differences between the operated and non-operated limb were observed during both evaluations (Fig. 6A). The same situation was observed for the total work measurements (both evaluations, Fig. 7A), and time to peak torque during knee extension at 30°/s velocity at the 24 months follow-up (p=0.0263) and at 60°/s velocity at the 12 months follow-up (p=0.0260). Importantly, there were no statistically significant differences in any of the isokinetics tests performed on the operated limbs, between the partial and the total Achilles tear subgroups. A significant deterioration was observed for the operated limbs in the total tear subgroup in multiple isokinetic parametes (Table 7, Fig. 5B, 6B and 7B).

**Table 7.**
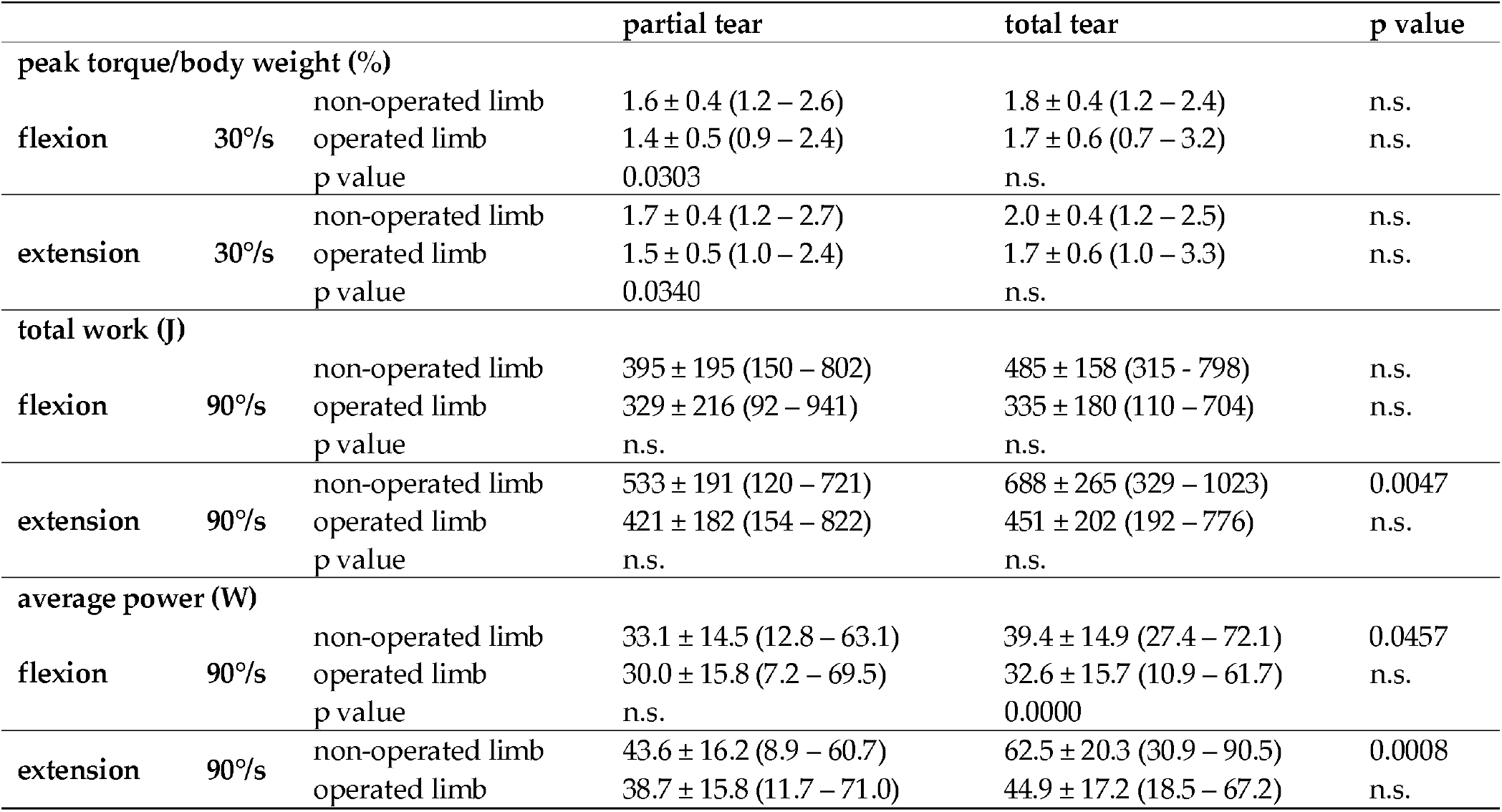
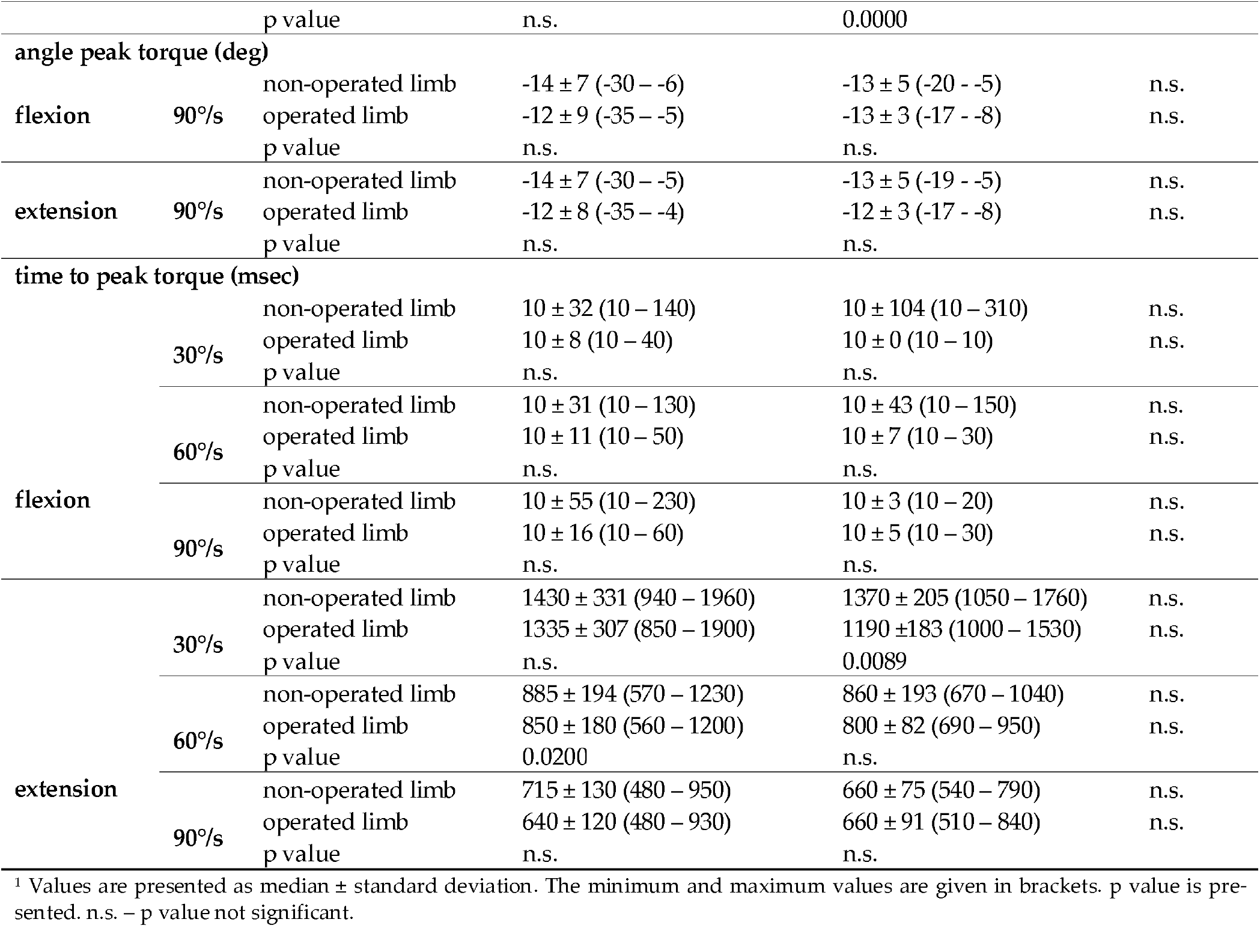
The comparison of the isokinetic outcomes of the surgical procedure between two Achilles tear subgroups.

**Figure 5.**
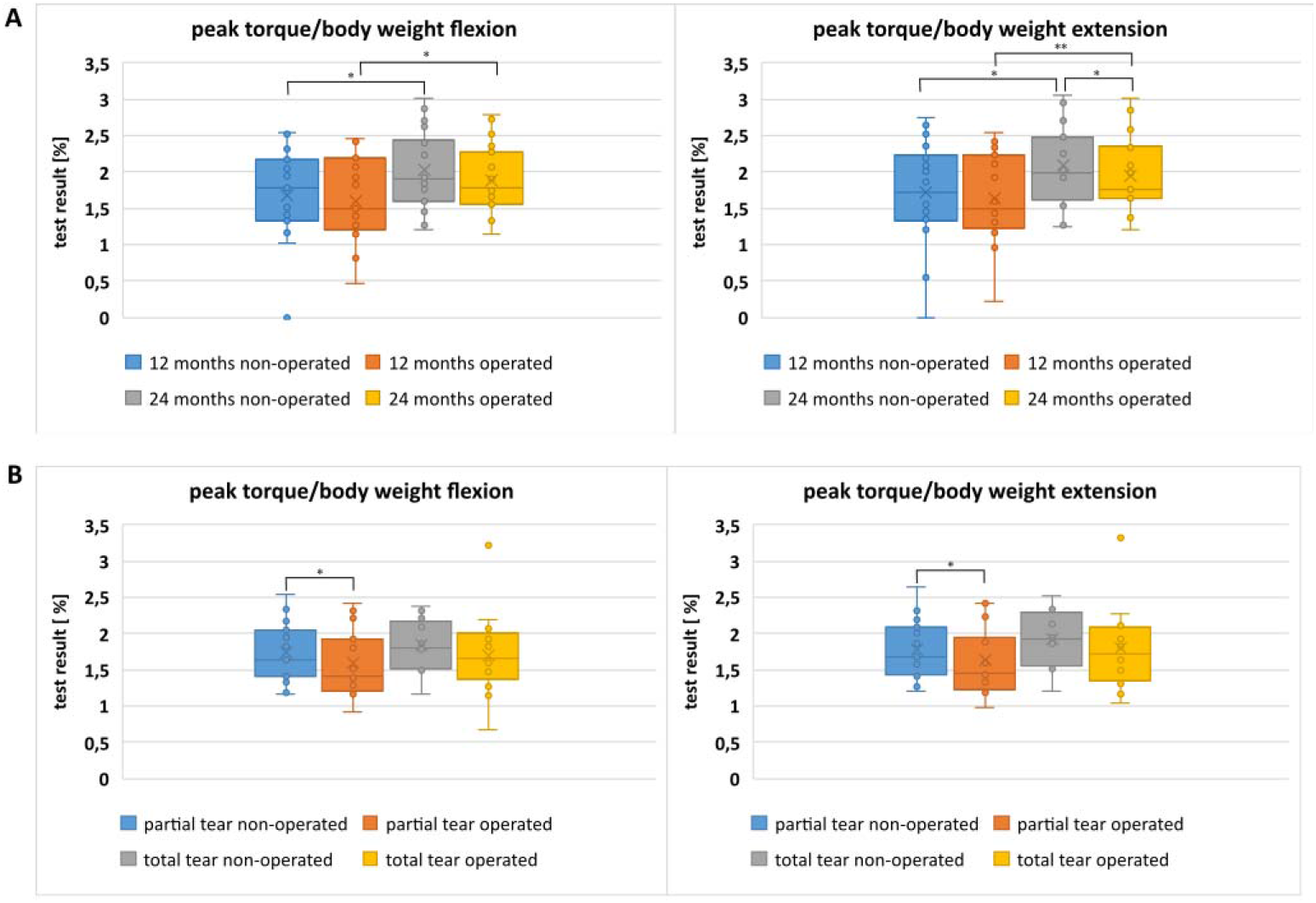
Evaluation of isokinetic differences between of the peak torque parameter during flexion and extension of operated and non-operated limbs (**A**) at 12 and 24 months postoperatively; (**B**) in two subgroups of partial and total Achilles tendon tear.

**Figure 6.**
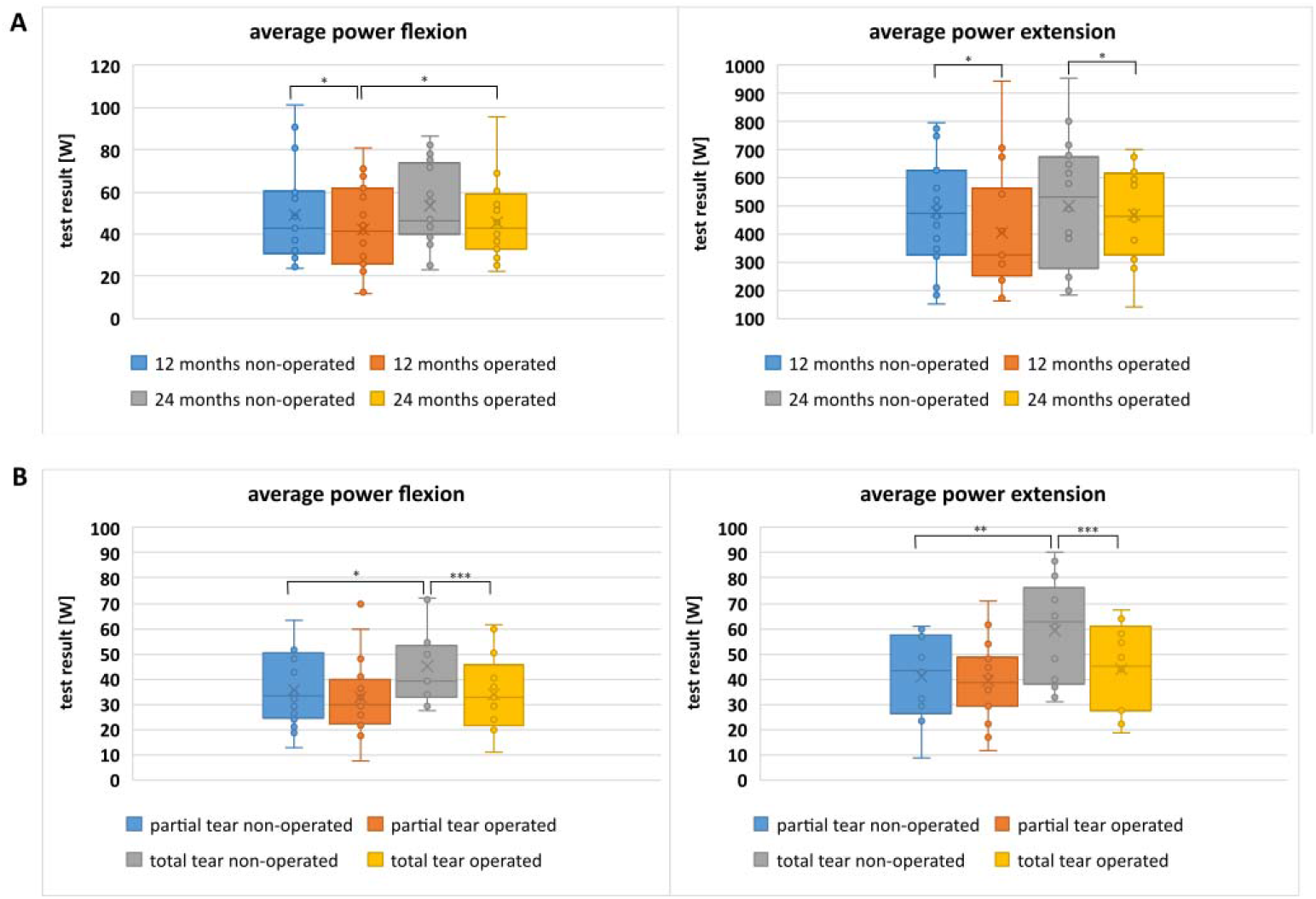
Evaluation of isokinetic differences between of the average power parameter during flexion and extension of operated and non-operated limbs (**A**) at 12 and 24 months postoperatively; (**B**) in two subgroups of partial and total Achilles tendon tear.

**Figure 7.**
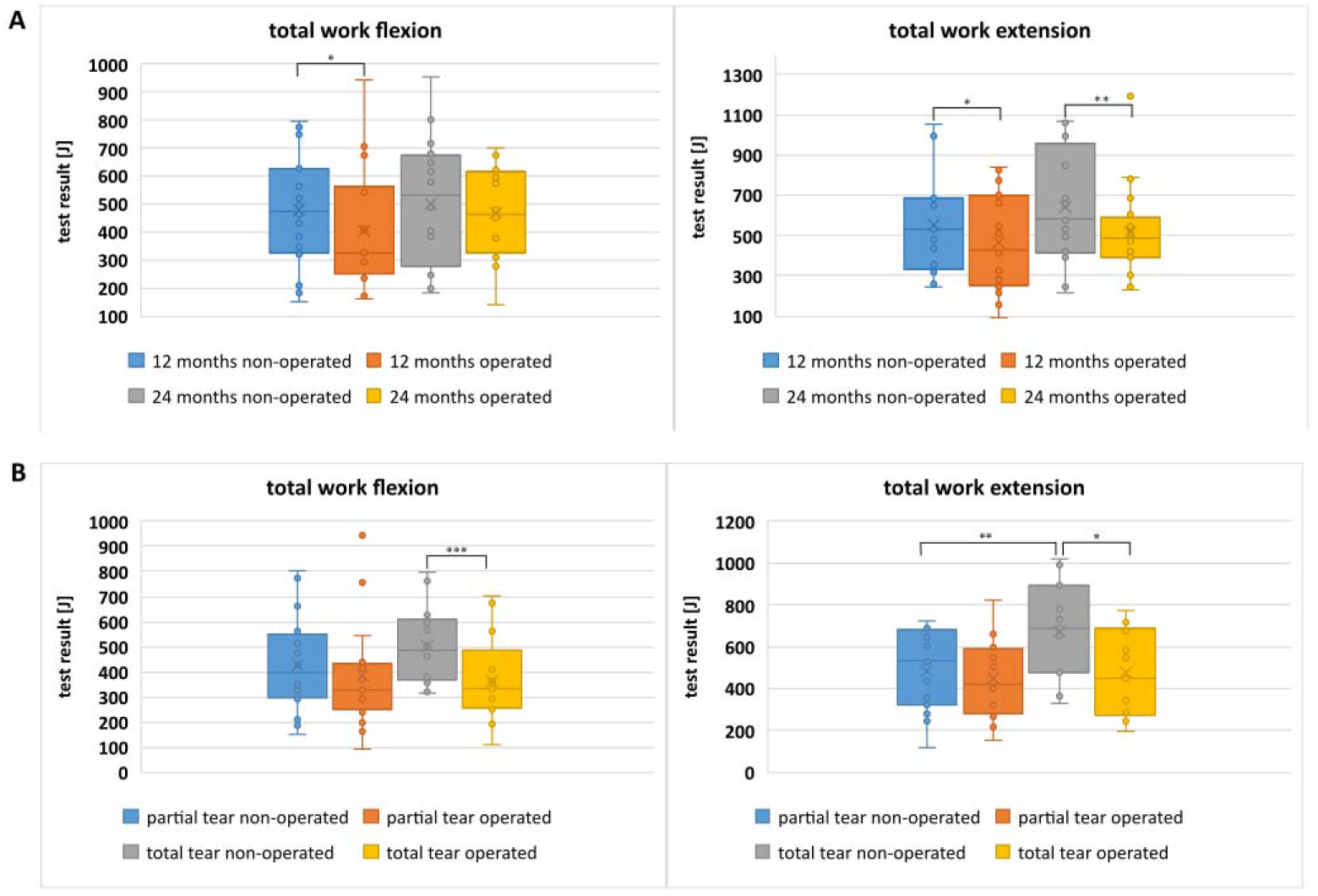
Evaluation of isokinetic differences between of the total work parameter during flexion and extension of operated and non-operated limbs (**A**) at 12 and 24 months postoperatively; (**B**) in two subgroups of partial and total Achilles tendon tear.

### Correlationd between the subjective and objective tests

Importantly, there were two strong correlations between the subjective and the objective tests. The VAS score (pain assesment) results highly correlated with the heel rise endurance test (r=-0.5457) and the results of the EQ-5D-5L score strongly correlated with the average power of the muscles durinf knee flexion (r=-0.5306).

## Discussion

The most important finding of our work is that the minimally invasive Achilles tendon reconstruction using semitendinosus and gracilis tendons with Endobutton stabilization enables satisfactory restoration of strength and function of the operated area within 24 months, as well as the overall satisfaction of patients. This allows patients to safely return to the daily physical activities.

The results presented in our study are in a good agreement with already published ones. Several authors have already tackled this point and presented restoring a full function of the operated Achilles tendon and the overall patient satisfaction. However, to our knowledge, this is the first study, which presents a detailed, versatile clinical, functional and isokinetic assessment of patients after Achilles tendon reconstruction. Multiple assessments, which described the outcomes of the Achilles reconstruction were based on two most popular subjective, patient-reported scales: Achilles tendon Total Rupture Score (ATRS) and American Orthopedic Foot and Ankle Society Score (AOFAS), combined with isokinetic tests or functional tests [16-20]. However, in these studies, a clear contradiction was observed between the subjective and the objective outcomes, which, in our opinion, might be the result of an insufficient number of the Achilles tendon parameters tested. For example, in 2014, Zhao et al. observed that some of the isokinetic parameters were significantly reduced in the operated limb [21]. Nevertheless, the patients did not report any deficits in function and were satisfied with the return to the physical fitness, despite clear deficits in the isokinetic and clinical assessment. In another study, two isolated isokinetic parameters were evaluated (peak torque of the plantar and dorsal flexor) and the authors observed two contradictory results for two knee positions [17]. All the above results indicate the need for a multifaceted approach to the Achilles tendon evaluations.

In the present study, the overall satisfactory outcomes are the resultant of multiple parameters: (i) equalization of the calf circumference between operated and non-operated limb; (ii) a statistically significant improvement in the subjective scores (ATRS, VAS and EQ-5D-L); (iii) better performance in functional tests (weight bearing lunge test, heel rise test, single leg hop); (iv) a statistically significant improvement in the isometric strength of the operated limb, measured in “away” motion; and (v) a statistically significant improvement in multiple lower limb muscles strength and endurance isokinetic parameters. All these results made us to the conclusion that the Achilles tendon function have been properly restored 24 months after the surgery. Moreover, a detailed examination of the partial and the total tear subgroups led us to the conclusion, that the minimally invasive Achilles tendon reconstruction using hamstring grafts enables satisfactory restoration of strength and function, despite the severeness of the damage.

## Data Availability

The datasets used and analysed during the current study are available from the corresponding author on reasonable request.

## Conclusions

The conclusion of our research is the confirmation of the high levels of satisfaction and function with the minimally invasive Achilles tendon reconstruction using hamstring grafts, the absence of the functional limitations, and the return to pre-injury activity, regardless the severeness of the initial Achilles damage.

## Author Contributions

Conceptualization, B.K. and T.P.; investigation, B.K, P.B., P.C., ł.S., J.K. and M.M.; data curation, B.K; writing—original draft preparation, B.K.; writing—review and editing, P.B.; supervision, P.B. and T.P. All authors have read and agreed to the published version of the manuscript.

## Institutional Review Board Statement

The study was conducted in accordance with the Declaration of Helsinki, and approved by the Bioethic Committee of Karol Marcinkowski Medical University in Poznań, by resolution no. 743/20.

## Informed Consent Statement

Informed consent was obtained from all subjects involved in the study.

## Acknowledgments

In this section, you can acknowledge any support given which is not covered by the author contribution or funding sections. This may include administrative and technical support, or donations in kind (e.g., materials used for experiments).

